# Integrative Multi-omics Approach Reveals the Molecular Characterization and Differences of ECM-PI3K-Akt Pathway among Coronary Artery Bypass Grafting Conduits with Clinical Implications

**DOI:** 10.1101/2024.08.11.24311581

**Authors:** Hai-Tao Hou, Huan-Xin Chen, Zheng-Qing Wang, Lei Xi, Jun Wang, Qin Yang, Guo-Wei He

## Abstract

**BACKGROUND:** A key problem for results of coronary artery bypass grafting (CABG) is different long-term patency of grafts (internal mammary artery [IMA], radial artery [RA], and saphenous vein [SV]).

**METHODS AND RESULTS:** We investigated the biological differences among IMA-SV, RA-SV, and IMA-RA using multi-omics approaches in order to explore new therapeutic targets. Trios of the human IMA, RA, and SV (n=72) from the CABG patients were studied using transcriptomics and proteomics. Differential mRNAs/proteins were validated by multiple reaction monitoring and real-time quantitative PCR in samples from new cohort of patients. Differentially expressed (DE) RNAs (60 mRNAs, 4 lncRNAs, 2 circRNAs) and 8 proteins in all three comparisons were identified. DE mRNAs and proteins were classified into 4 correlations (non-DE RNAs/non-DEPs, DE RNAs/non-DEPs, non-DE RNAs/DEPs, and DE RNAs/DEPs). Eleven correlated DE mRNAs/DEPs (TSP1, TENA, TENX, VTNC, LAMA4, CO6A3, COMP, ITA1, DAG1, ITA5, and ITA8) were found in ECM-PI3K-Akt pathway, which may play important roles in vasodilation, stenosis, angiogenesis, platelet activation, inflammation, ECM remolding, and atherosclerosis. Importantly, lower TSP1 in IMA or RA than that in SV, lower TENA and LAMA4 in IMA than that in SV or RA, and higher ITA8 in IMA than that in RA may be the reasons of different long-term patency.

**CONCLUSIONS:** ECM-PI3K-Akt pathway with DE mRNAs and proteins may be the major pathway related to the differences among three grafting vessels. This study provides new insights into the biological differences of the grafts and may form new therapeutic targets for improving the long-term results of CABG.

**Clinical Perspective:** **What Is New?**

- We presented a human vessel-specific map on both RNA patterns and protein profiling in three major coronary artery bypass grafting (CABG) grafts: internal mammary artery (IMA), radial artery (RA), and saphenous vein (SV). DE mRNAs and proteins were classified into 4 correlations (non-DE RNAs/non-DEPs, DE RNAs/non-DEPs, non-DE RNAs/DEPs, and DE RNAs/DEPs).
- We revealed that ECM-PI3K-Akt pathway is the major pathway related to the differences among three major CABG grafting vessels including abundant differentially expressed mRNAs and proteins (TSP1, TENA, TENX, VTNC, LAMA4, CO6A3, COMP, ITA1, DAG1, ITA5, and ITA8).
- We also revealed that 12 correlated mRNAs and proteins (SUSD5, CO8A1, 3HAO, SRBS2, AIF1L, EFHD1, DESM, TSP1, POSTN, TGM2, HMCN2, and CO6A3) had differences between the arteries and the vein. Five correlated mRNAs and proteins (SUSD2, COCA1, AL1A1, ITA8, and ITIH1) had differences only in IMA-RA.
- Lower TSP1 in IMA or RA than that in SV, lower TENA and LAMA4 in IMA than that in SV or RA, and higher ITA8 in IMA than that in RA may be the reasons of different long-term patency.

**What Are the Clinical Implications?**

- This study reveals that the ECM-PI3K-Akt pathway is the major pathway related to the differences among three major CABG grafting vessels including abundant differentially expressed mRNAs and proteins and that the differences in this signaling pathway likely account for the differences in the long-term patency. Therefore, the study provides scientific evidence for why the grafts have different long-term patency at the biological basis in CABG.
- The study provides new insights into the new therapeutic targets for improving the results of CABG.

---

The grafting conduits used in coronary artery bypass grafting (CABG) surgery are autologous arteries (internal mammary artery [IMA] and radial artery [RA]) and veins (saphenous vein [SV]). Long-term patency of these conduit vessels used as grafts is different even under the same surgical technique and drug treatment. As a recent study reveals, the 5 year patency was 97.7% for IMA, 90.6% for RA, and 82.5% for SV^1^. In fact, the patency for 10 years or longer was 85-95% for IMA, 89-91% for RA, and 50-60% for SV^2^ (**Table 1**). In addition, graft-related complications represent the most common cause of perioperative myocardial ischemia, accounting for approximately 2/3 of cases^3^.

**Table 1.**
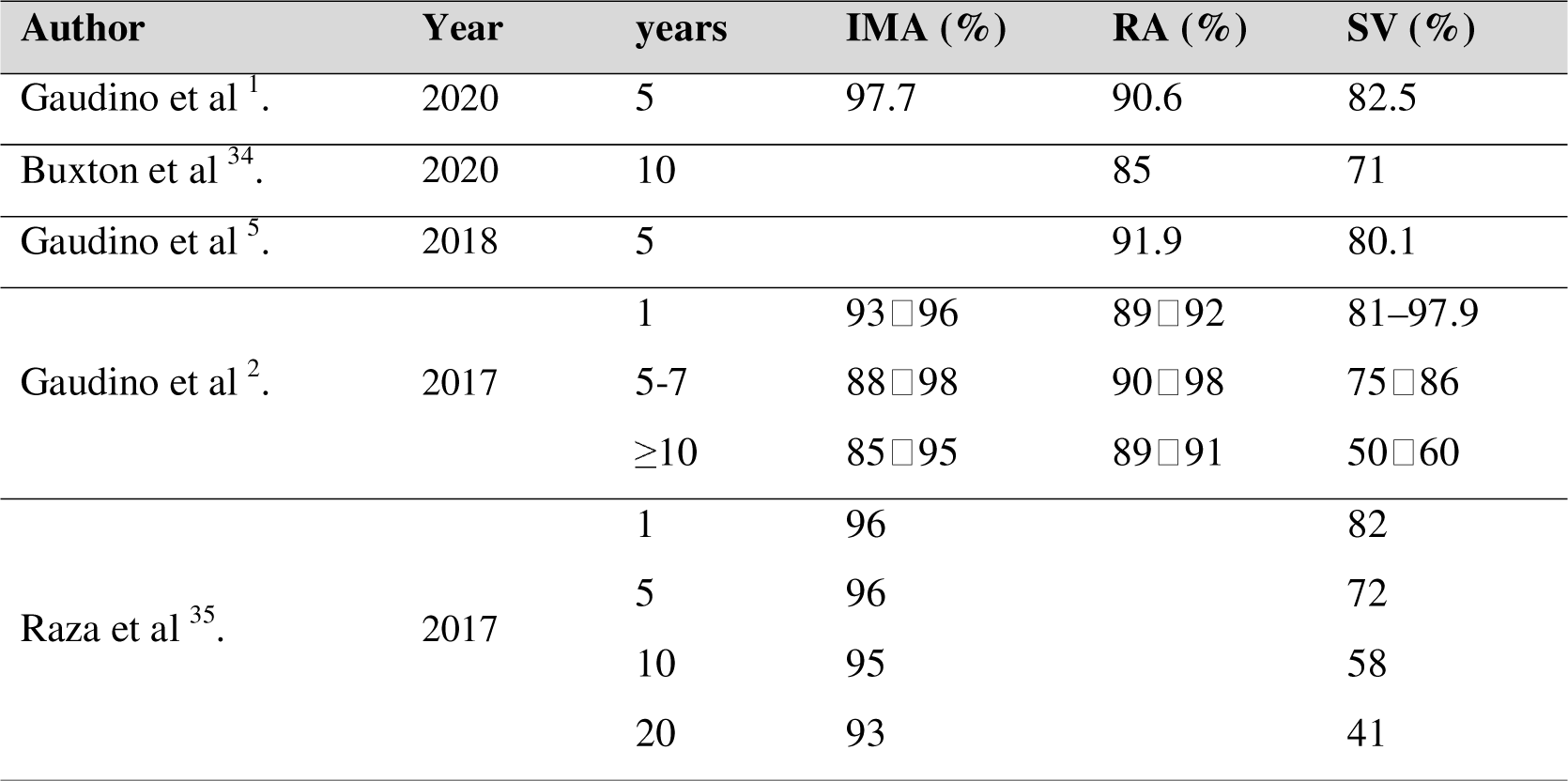
The patency rates of internal mammary artery (IMA), radial artery (RA), and saphenous vein (SV) used in coronary artery bypass grafting.

| Author | Year | years | IMA (%) | RA (%) | SV (%) |
| --- | --- | --- | --- | --- | --- |
| Gaudino et al <sup>1</sup>. | 2020 | 5 | 97.7 | 90.6 | 82.5 |
| Buxton et al <sup>34</sup>. | 2020 | 10 |  | 85 | 71 |
| Gaudino et al <sup>5</sup>. | 2018 | 5 |  | 91.9 | 80.1 |
| Gaudino et al <sup>2</sup>. | 2017 | 1 | 93–96 | 89–92 | 81–97.9 |
|  |  | 5-7 | 88–98 | 90–98 | 75–86 |
|  |  | ≥10 | 85–95 | 89–91 | 50–60 |
| Raza et al <sup>35</sup>. | 2017 | 1 | 96 |  | 82 |
|  |  | 5 | 96 |  | 72 |
|  |  | 10 | 95 |  | 58 |
|  |  | 20 | 93 |  | 41 |

The left IMA is usually the first choice for left anterior descending artery. As to the second best graft, a meta-analysis demonstrated an angiographic superiority of RIMA and RA over SVG^4^. Vein grafts are also the common choice even though arteries have higher patency^2, 5^ due to the fact that more than three grafts are frequently needed in patients with severe coronary disease^6^.

Biological characteristics are related to the different long-term patency of the grafts^7^. Different biological mechanisms including thrombosis caused by endothelial dysfunction and atherosclerotic plaque formation are not conducive to the long-term patency^2, 8, 9^. Differences exist between the arterial and venous grafts^10^ and among the arteries used as conduits^7, 11, 12^. In particular, the basal release of nitric oxide (NO) and endothelium-derived hyperpolarizing factor are significantly greater in human IMA than in SV^10^ and RA^13^, reported by us. Others have also reported that SV has a greater propensity for intimal hyperplasia than arterial grafts, but RA behaves similarly to IMA^14^.

It is well known that the development processes of arteries and veins are fundamentally different^15^. However, systematic studies at the transcriptomics and proteomics levels to analyze the differences among arterial and venous grafts have not been reported, although omics technologies are widely used^16–19^.

The present study aimed to test the hypothesis that there are intrinsic differences in biological characteristics among the CABG grafts at the mRNA and protein levels and the differences may involve particular cellular signaling pathways. In this study, we used integrated transcriptomics and proteomics to systematically investigate the differences among the IMA, RA, and SV at the RNA and protein level in order to reveal the intrinsic differences in biological characteristics among the CABG grafts and possibly involved signaling pathway(s).

## METHODS

### Preparation of human grafts

Trios of discarded vascular segments (n=72) from 24 patients simultaneously receiving these three grafts during first-time CABG without other concurrent operations were studied to reveal the similarities and differences among the three conduits (IMA, RA, and SV) without obvious vascular diseases and between IMA and SV (IMA-SV), RA and SV (RA-SV), or IMA and RA (IMA-RA). The experimental design of the study is depicted in **Fig. 1**.

**Figure 1.**
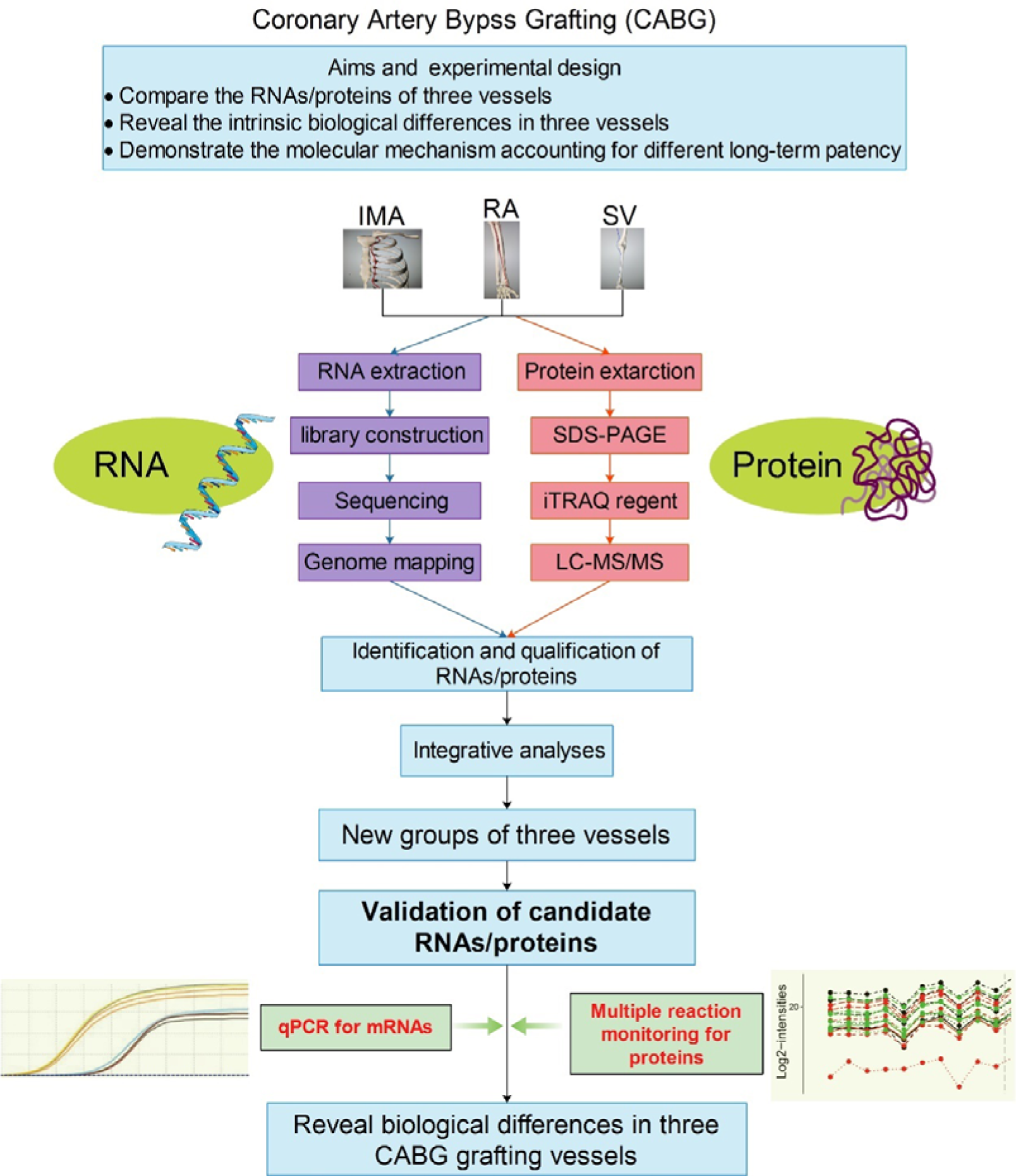
The experimental design. Internal mammary artery (IMA), radial artery (RA), and saphenous vein (SV) segments from coronary artery bypass grafting (CABG) were studied using transcriptomics and proteomics, followed by validation in samples from a new cohort of patients. Note that the results of omics studies were validated by qPCR for mRNAs and multiple reaction monitoring for proteins.

### RNA-sequencing (RNA-Seq) and liquid chromatography-tandem mass spectrometry (LC-MS/MS)

Total RNA was extracted and sequencing was conducted using DESeq2 using HiSeq X Ten platform. Differential expression analysis was performed by using DESeq2. Total protein was extracted and mass spectra were analyzed using LC-MS/MS.

### Criterias for differential screening

The differential RNAs or proteins were defined as follows: (1) *q* values (*P*-adjusted values) < 0.05; (2) fold change was more than 2 or less than 0.5 in transcriptomics and more than 1.2 or less than 0.83 in proteomics; (3) in validation of proteins by using MRM the fold change was set at 1.5 and 0.67.

### Bioinformatics analysis and validation

Three major Gene Ontology (GO) categories (biological process, cellular component, and molecular function), Kyoto Encyclopedia of Genes and Genomes (KEGG) enrichment, and protein-protein interaction (PPI) of annotated different expression RNAs and proteins were analyzed. Relative expression of mRNA was validated by qPCR method in 30 grafts. MRM analyses were performed using QTRAP 6500 mass spectrometer in another 30 grafts.

### Statistical analysis

Hierarchical clustering was used to demonstrate the relationships using differentially expressed (DE) RNAs and differentially expressed proteins (DEPs). The comparison between two groups was conducted with Student’s t test. *P* < .05 was considered significant. Data were analyzed using SPSS 20.0 software and GraphPad Prism. Data were represented as mean ± SEM.

## RESULTS

### RNAs expression analysis and heterogeneity in grafts

After mapping sequenced reads to reference genome, we finally identified 71,991, 73,593, and 70,664 RNAs (mRNA, lncRNA, and circRNA), in IMA, RA, and SV respectively (**Fig. 2A**). Further, **Fig. 2B-D** show the overlap of mRNA, lncRNA, and circRNA respectively. **Fig. 2E-G** show continuous declines with small fluctuations of all the chromosomes. **Fig. 2H-K** summarize the number of three types of RNA in three grafts according to the Fragments Per Kilobase Million (FPKM) value. The most abundant RNAs were summarized in IMA-SV (*NEMF*, et al. **Fig. 2L**), RA-SV (LOC107987206, et al. **Fig. 2M**), and (circ_0006940, et al. **Fig. 2N**). **Fig. 2O-Q** show the general differences among IMA, RA, and SV.

**Figure 2.**
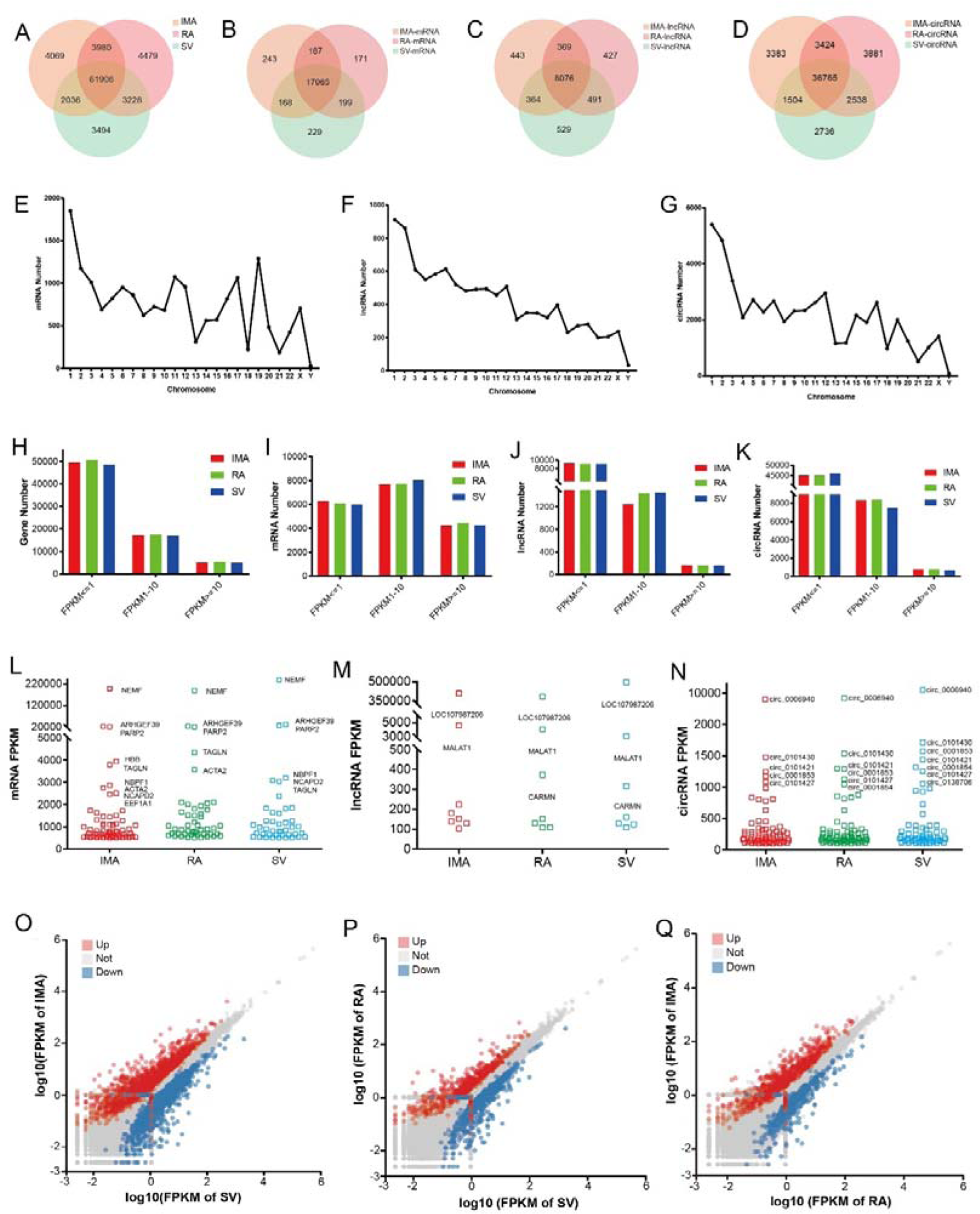
Transcriptome analyses of internal mammary artery (IMA), radial artery (RA), and saphenous vein (SV). **A-D**, venn diagram showing the number of total RNAs, mRNAs, lncRNAs, and circRNAs in each graft. **E-G**, number of mRNAs, lncRNAs, and circRNAs in all chromosomes. **H-K**, number of total RNAs, mRNAs, lncRNAs, and circRNAs in each graft based on transcript abundance (FPKM value). **L-N**, the most abundant mRNAs (FPKM > 500), lncRNAs (FPKM > 100), and circRNAs (FPKM > 100) in each graft. **O-Q**, pairwise comparisons showing RNAs expression in each graft according to FPKM values on a log10. Red color represents that the FPKM of Y axis is significantly more than that of X axis. In opposite, blue color represents that the FPKM of X axis is more than that of Y axis. Gray color represents that there is no difference.

According to the expression abundance (FPKM) in this study, we found that: (1) *GJA5*, *GJA3*, *PCK1*, *FBP2*, and *ITLN1* in arteries were more abundant than that in veins; (2) *CCL7*, *MYH10*, *S100A9*, and *VMP1* in IMA were more abundant than that in RA or SV; (3) *MSX2* in RA was more abundant than that in IMA or SV; 4) *HOXC10*, *HOXC11*, *HOXC13*, *HMCN2*, *PRAC2*, *KRTAP1-5*, and *NRP2* in SV were more abundant than that in IMA or RA; (4) *DES* was significantly different among the three comparisons.

Further, DE RNAs were identified between IMA-SV (3,239 DE RNAs [1,777 mRNAs, 462 lncRNAs, and 1,000 circRNAs, **Tables S1-3**], between RA-SV (1,246 [788 mRNAs, 152 lncRNAs, and 306 circRNAs, **Tables S4-6**], and between IMA-RA (1,226 [739 mRNAs, 220 lncRNAs, and 267 circRNAs, **Tables S7-9**], respectively. **Figure 3A-C** show the overlap of DE mRNA, lncRNA, and circRNA in different comparison groups. Total of 66 RNAs (60 mRNAs, 4 lncRNAs, and 2 circRNAs) had differences in all the three comparisons by venn diagram with connections about 11 mRNAs in the 60 mRNAs (**Fig. 3G**) by protein-protein (RNAs products) interaction analysis. KEGG network diagram (**Fig. 3H**) shows that 14 mRNAs (such as *PDGFB*) involved into 10 KEGG pathways. **Fig. 3I** shows top 20 significantly enriched different KEGG pathways (*P* < .05) using 66 co-DE RNAs in all the three comparisons.

**Figure 3.**
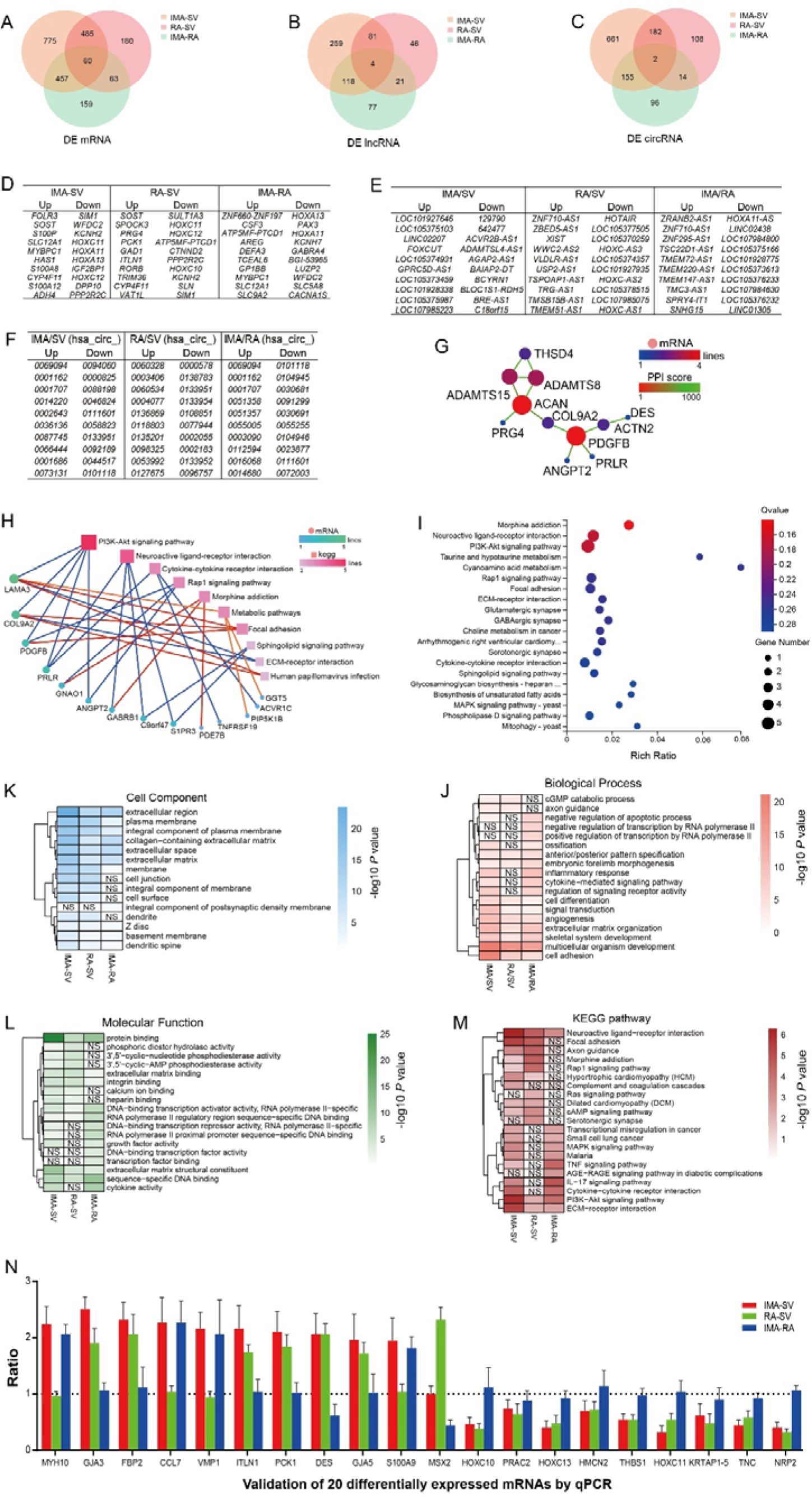
Differentially expressed RNAs in IMA, RA, and SV. **A-C**, venn diagram showing the overlap of differentially expressed mRNAs, lncRNAs, and circRNAs in each comparison between two grafts among IMA, RA, and SV. **D-F**, top 10 up-regulated and 10 down-regulated mRNAs, lncRNAs, and circRNAs in each comparison between two grafts. **G**, protein-protein (products of 60 co-differentially expressed mRNAs) interaction analysis. **H**, KEGG network diagram of the 60 co-differentially expressed mRNAs. Fourteen mRNAs are involved in 10 KEGG pathways. **I**, top 20 statistically significantly different KEGG pathways using 60 co-differentially expressed RNAs. **J-M**, Hierarchical clustering of the RNA profiles revealing striking differences in biological process, cell component, molecular function, and KEGG pathways based on -log10 of P value among 3 comparisons of each two grafts. **N**, 20 differentially expressed mRNAs validated by qPCR method. Also see Tables S1-9 in the Supplemental Material for the details. NS, not significant.

Hierarchical clustering of the RNA profiles revealed striking differences about GO and KEGG pathways. For biological process (**Fig. 3J**), there was the most significant difference about multicellular organism development in all the three comparisons. Similarly, the differences in cellular component (**Fig. 3K**), molecular function (**Fig. 3L**), and KEGG pathways (**Fig. 3M**) are also demonstrated. Twenty DE mRNAs were validated (**Fig. 3N**) and the primer sequences of each mRNA synthesized in Sangon Biotech were listed in **Table S10**.

### Protein expression analysis

In proteomic studies, total of 124,244 spectra, 12,547 peptides, 12,065 unique peptides, corresponding to 3,096 proteins from the LC-MS/MS analysis were identified (**Fig. 4A**). Of these proteins, 4.5% (138 of 3,096, 56 up-regulated and 82 down-regulated, **Table S11**) in IMA-SV, 3.1% (95 of 3,096, 41 up-regulated and 54 down-regulated, **Table S12**) in RA-SV, and 2.3% (71 of 3,096, 31 up-regulated and 40 down-regulated, **Table S13**) in IMA-RA were screened as DEPs. **Fig. 4B** shows the top 10 up-regulated and down-regulated DEPs respectively.

**Figure 4.**
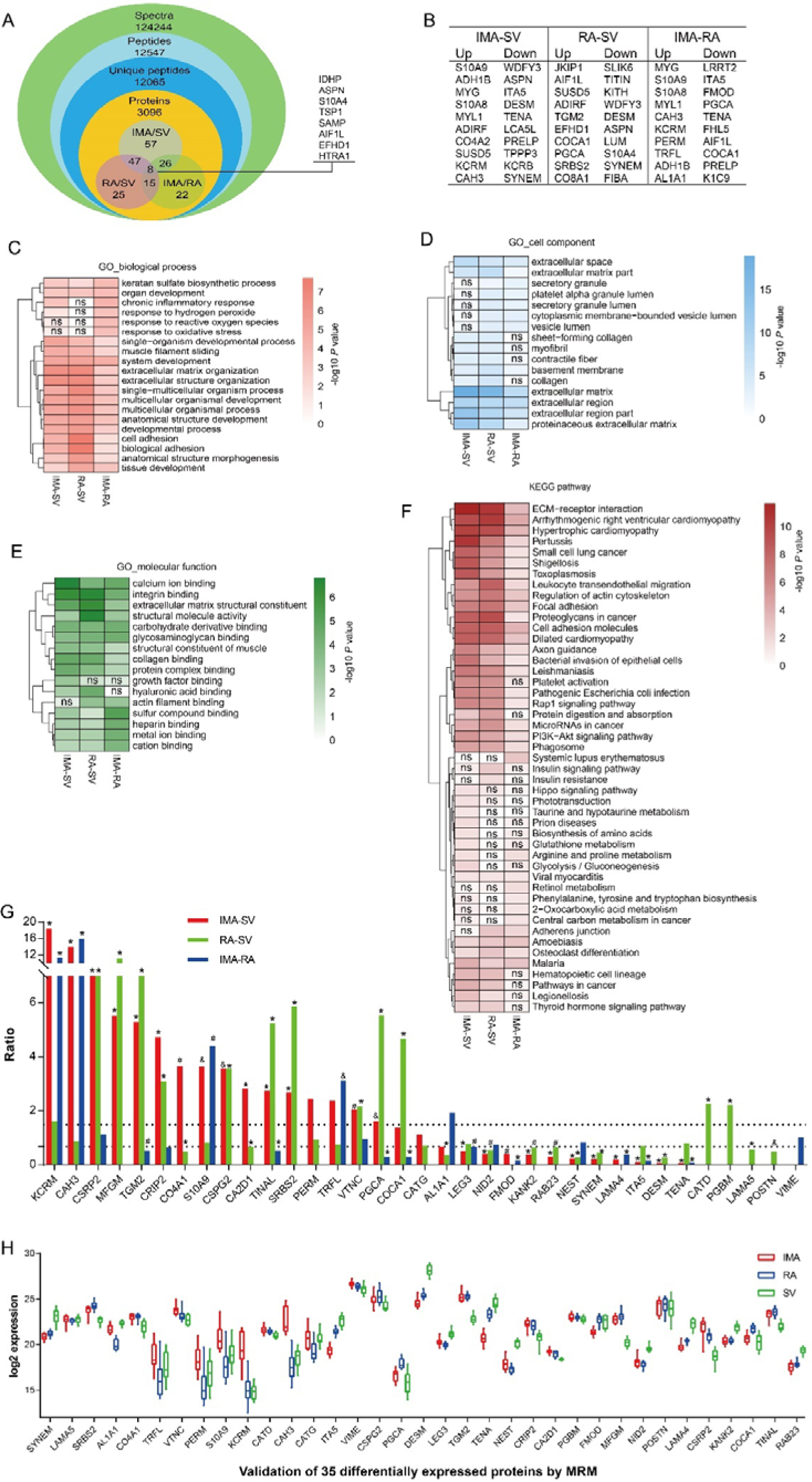
Differentially expressed proteins in IMA, RA, and SV - proteomics analyses. **A**, venn diagram showing the overlap of spectra, peptides, unique peptides, proteins, and differentially expressed proteins based on the comparison between each two grafts among IMA, RA, and SV. **B**, top 10 up-regulated and 10 down-regulated proteins in the comparison between each two grafts. **C-F**, heat map displaying the significantly different GO categories (biological process, cell component, and molecular function) and KEGG pathways in the comparison between each two grafts. **G**, successful verification of 35 differentially expressed proteins in the comparison between each two grafts using Multiple Reaction Monitoring method in a new cohort of patients. The percentage of the accordance between the verification in the new cohort and the omics results regarding the 35 differentially expressed proteins is 85.7% in IMA-SV, 91.4% in RA-SV, and 57.1% in IMA-RA, respectively. The order of proteins is arranged according to the value of IMA-SV. **H**, Details of the log2 expression of all the 35 validated proteins. Also see Tables S11-13 in the Supplemental Material for the details. IMA, internal mammary artery. RA, radial artery. SV, saphenous vein. MRM: Multiple Reaction Monitoring. NS, not significant. ^&^P < 0.05, ^#^P < 0.01, *P < 0.001.

Top 10 significant GO terms were put together to compare the significance among three comparisons (IMA-SV, RA-SV, and IMA-RA) (**Fig. 4C-E**). Similarly, the comparisons on KEGG were performed with all significant pathways (**Fig. 4F**). Regarding biological process (**Fig. 4C**), there were significant differences in response to reactive oxygen species, oxidative stress only in IMA-RA. Similarly, the difference in cellular component (**Fig. 4D**), molecular function (**Fig. 4E**), and KEGG (**Fig. 4F**) was also shown. In addition, in pathways such as arginine and proline metabolism, glycolysis/gluconeogenesis, biosynthesis amino acids, and glutathione metabolism pathways, the most significant differences were seen in IMA-SV and IMA-RA, but not in RA-SV (**Fig. 4F**).

Thirty-five proteins were chosen for further expression validation by MRM technology (**Fig. 4G**). The percentages of successful MRM validation are 85.7% (30/35), 91.4% (32/35), and 57.1% (20/35) in IMA-SV, RA-SV, and IMA-RA respectively. **Fig. 4H** shows log2 expression of these 35 proteins in 3 grafts respectively.

### Comparisons between the arteries and the vein

There were significant differences in 485 mRNAs (**Fig. 3A**), 81 lncRNAs (**Fig. 3B**), 182 circRNAs (**Fig. 3C**), and 47 proteins (**Fig. 4A**) between the artery (IMA or RA) and the SV. Pathways analysis (using all the DEPs) showed that up to 28 pathways (such as ECM-receptor and platelet activation) were more different between the arteries and the vein (IMA-SV and RA-SV) than between the arteries (IMA-RA) (**Fig. 4F**).

### Comparisons between the two arteries

There were significant differences in 159 mRNAs (**Fig. 3A**), 77 lncRNAs (**Fig. 3B**), 96 circRNAs (**Fig. 3C**), and 22 proteins (**Fig. 4A**) between the two arteries (IMA-RA). Pathways analysis (using all the DEPs) showed that 6 pathways (such as Retinol metabolism) were more significant between the arteries (IMA-RA) than between the arteries and the vein (IMA-SV or RA-SV) (**Fig. 4F**).

### Comparison of IMA to other two grafts (RA or SV)

The IMA has significant differences compared to the other two grafts. There were significant differences in 457 mRNAs, 179 lncRNAs, 155 circRNAs, and 26 proteins between the IMA and the other two grafts (IMA-SV or IMA-RA). Pathways analysis (using all the DEPs) showed that 10 pathways (such as arginine and proline metabolism) were more significant between the IMA and the other two grafts (IMA-SV or IMA-RA) (**Fig. 4F**). These findings may reveal the most important reason why the IMA has better long-term patency than the other two grafts.

### Comparisons among three grafts

Among the three grafts, there were significant differences in 60 mRNAs (**Fig. 3A**), 4 lncRNAs (*GPRC5D-AS1*, *LOC101927646*, *LOC105373479*, and *DLX6-AS1*) (**Fig. 3B**), 2 circRNAs (*hsa_circ_0101406* and *hsa_circ_0058840*) (**Fig. 3C**) and 8 DEPs (IDHP, ASPN, S10A4, TSP1, SAMP, AIF1L, EFHD1, and HTRA1) (**Fig. 4A**). Interaction analyses indicated that 11 mRNAs (*PDGFB*, *PRLR*, *ACTN2*, *ANGPT2*, *COL9A2*, *DES*, *ACAN*, *PRG4*, *ADAMTS8*, *ADAMTS15*, and *THSD4*) (**Fig. 3G**) are related to proatherogenic process.

### Comparisons of correlated mRNAs and proteins among three grafts

Comprehensive correlation analyses showed that more than 93% proteins could be mapped to RNAs. The correlation between the RNAs and the coded proteins was categorized at 4 levels, i.e., non-DE RNAs/non-DEPs, DE RNAs/non-DEPs, non-DE RNAs/DEPs, and DE RNAs/DEPs (**Fig. 5A**)

**Figure 5.**
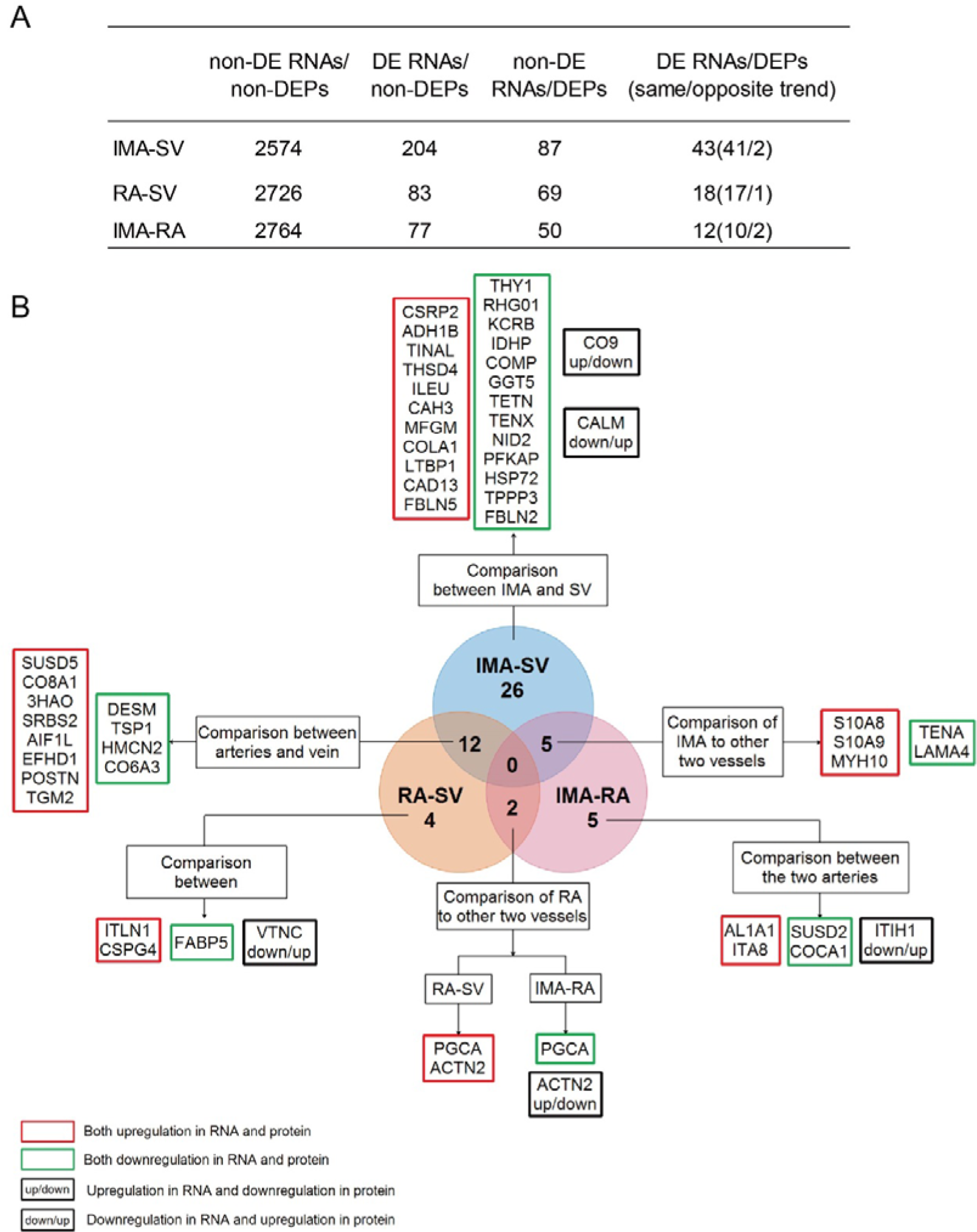
Correlation analysis between DE RNAs and DEPs. A, summary of four different levels of the correlations (non-DE RNAs/non-DEPs, DE RNAs/non-DEPs, non-DE RNAs/DEPs, and DE RNAs/DEPs). B, six kinds of comparisons with DE RNAs/DEPs to illustrate the differences. Red box: upregulation in both RNA and protein; green box: downregulation in both RNA and protein; black box: opposite regulation in both RNA and protein. DE, differentially expressed. DEPs, differentially expressed proteins. IMA, internal mammary artery. RA, radial artery. SV, saphenous vein. Also see Tables S14-16 in the Supplemental Material for the details.

In IMA-SV, we found 93.9% (2,908 of 3,096) proteins could be mapped to RNAs. Of the correlated mRNAs and proteins, 88.5% (2,574) were non-DE RNAs/non-DEPs; 10.0% (291) were DE RNAs/non-DEPs or non-DE RNAs/DEPs; 1.5% (43) were DE RNAs/DEPs.

In RA-SV group, 93.5% (2,896 of 3,096) proteins could be mapped to RNAs. Of the correlated mRNAs and proteins, 94.1% (2,726) were non-DE RNAs/non-DEPs; 5.2% (152) were DE RNAs/non-DEPs or non-DE RNAs/DEPs; 0.6% (18) were DE RNAs/DEPs.

In IMA-RA group, 93.8% (2,903 of 3,096) proteins could be mapped to RNAs. Of the correlated mRNAs and proteins, 95.2% (2,764) were non-DE RNAs/non-DEPs; 4.4% (127) were DE RNAs/non-DEPs or non-DE RNAs/DEPs; 0.4% (12) were DE RNAs/DEPs.

The details of the correlation in each group are shown in **Tables S14-16**.

Venn analysis revealed the specific DE RNAs and DEPs in the 3 comparisons. As to the differences between the arteries (IMA and RA) and the vein (SV), 12 correlated mRNAs and proteins (SUSD5, CO8A1, 3HAO, SRBS2, AIF1L, EFHD1, DESM, TSP1, POSTN, TGM2, HMCN2, and CO6A3) (**Fig. 5B**) were found different. Further, the IMA had differences in 5 correlated mRNAs and proteins (MYH10, S10A8, S10A9, TENA, and LAMA4), compared to the other two grafts (RA and SV) whereas the RA had differences in 2 correlated mRNAs and proteins (PGCA and ACTN2), compared to the other grafts (IMA and SV).

In addition, there were 5 correlations (SUSD2, COCA1, AL1A1, ITA8, and ITIH1) that were only different between IMA and RA and 26 correlations only different between IMA and SV. Further, there were 4 correlations (ITLN1, FABP5, CSPG4, VTNC) that were only different between RA and SV.

### Analyses of the DE RNAs and DEPs

Spearman correlation analysis (**Fig. 6A-C**) indicates strong relationship between differentially expressed RNAs and DEPs. The spearman values in IMA-SV, RA-SV, and IMA-RA were 0.91, 0.83, and 0.74, respectively. **Supplementary Figure S1** shows all the correlations of non-DE RNAs/non-DEPs, DE RNAs/non-DEPs, non-DE RNAs/DEPs, and DE RNAs/DEPs).

**Figure 6.**
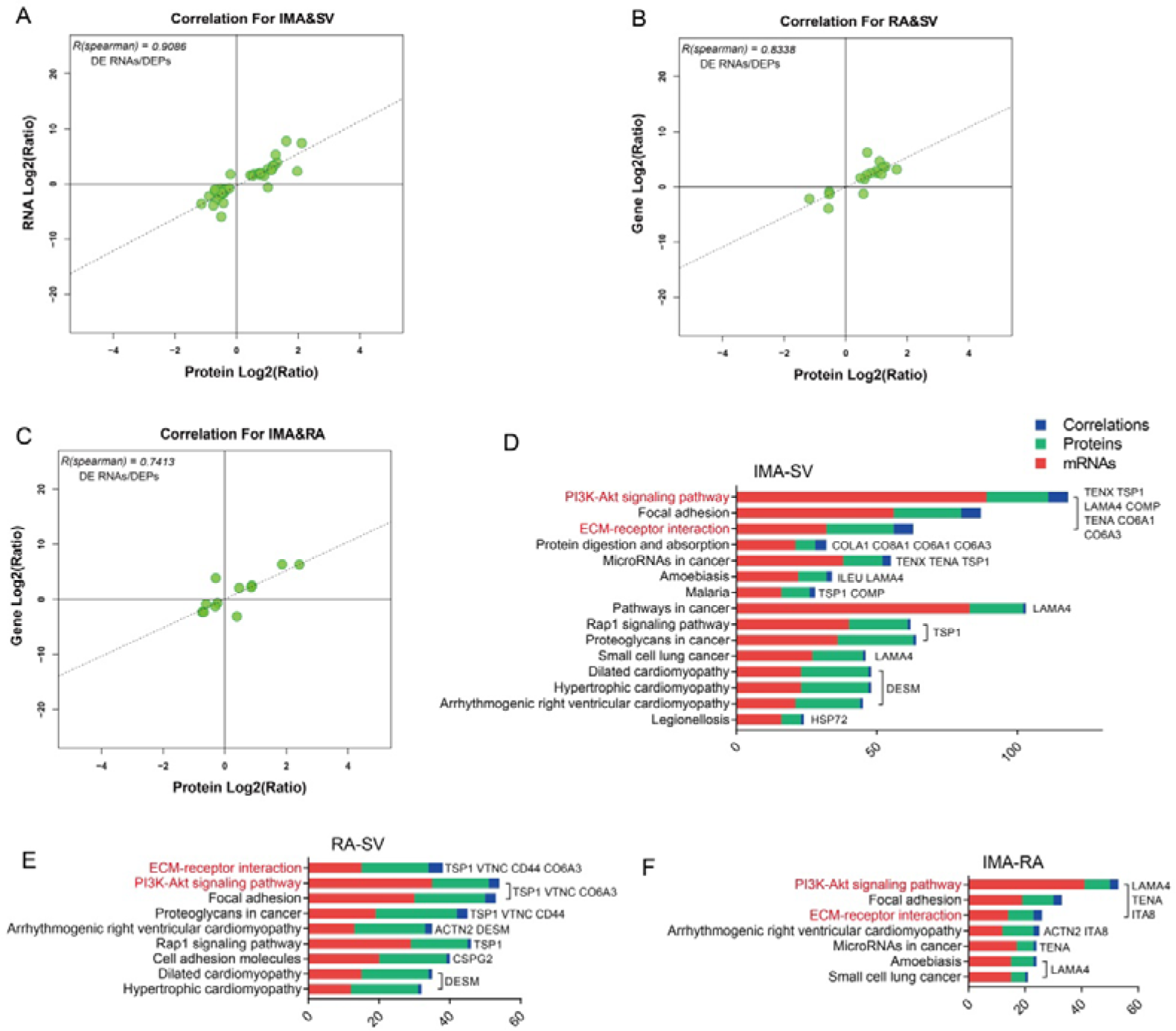
Spearman correlation and pathway analyses using DE RNAs and DEPs. , spearman values in IMA-SV, RA-SV, and IMA-RA are 0.91, 0.83, and 0.74, respectively. D-F, significantly enriched pathways (involved at least one correlation of DE RNAs and DEPs) in both transcriptomics and proteomics. The correlations involved in these pathways were listed on the right of the bar. ECM-PI3K-Akt pathway (red characters) included most correlated mRNAs and proteins in IMA-SV (7), RA-SV (3), and IMA-RA (3), respectively. DE, differentially expressed. DEPs, differentially expressed proteins. IMA, internal mammary artery. RA, radial artery. SV, saphenous vein.

**Fig. 6D-F** show the significantly enriched pathways (involved at least one correlation of DE RNAs and DEPs) in both transcriptomics and proteomics. In these pathways, ECM-PI3K-Akt pathway included most correlated DE mRNAs and proteins in IMA-SV (7), RA-SV (3), and IMA-RA (3), respectively. **Supplementary Figure S2** shows the enriched GO categories in biological process, cellular compartment, and molecular function.

### Patency of the grafts

All patients were followed up with average of 25.9 ± 5.7 months. Thirteen patients had coronary angiogram. The total grafts were 47. All 28 arterial grafts (13 IMA and 15 RA) were patent without stenosis. In contrast, in 19 SV grafts, there were 4 grafts that had significant stenosis (*P* = 0.022 compares to the arterial grafts, Fisher’s exact test; **Table S17**).

## Discussion

In this study we for the first time by using integrated transcriptomics and proteomics methods in vessels used as CABG grafts found that 1) there are a larger number of differentially expressed RNAs and proteins between the arteries and the vein and between the two arteries; 2) correlated mRNAs and proteins in 4 levels (non-DE RNAs/non-DEPs, DE RNAs/non-DEPs, non-DE RNAs/DEPs, and DE RNAs/DEPs) were clarified; 3) bioinformatics analyses including biological process, cell component, molecular function, and KEGG pathways were compared to reveal the differences among IMA-SV, RA-SV, and IMA-RA; 4) ECM-PI3K-Akt pathway is the major signaling pathway in the differences among these grafts.

### Importance of ECM-PI3K-Akt pathway and eNOS in development of stenosis, angiogenesis, and atherosclerosis

We found that enriched ECM-PI3K-Akt pathway is the most important signaling pathway with more DE RNAs and DEPs (TSP1, TENA, TENX, VTNC, LAMA4, CO6A3, COMP, ITA1, DAG1, ITA5, and ITA8) than other pathways. **Fig. 7** is a schema to illustrate a new concept that the interaction between ECM-PI3K-Akt and eNOS are involved in the long-term patency. All these DE mRNAs and proteins (**Fig. 5B**), particularly TSP1, TENA, and LAMA4, have certain connections with nitric oxide and play important roles in vasodilation, stenosis, angiogenesis, platelet activation, inflammation, ECM remolding, and atherosclerosis.

**Figure 7.**
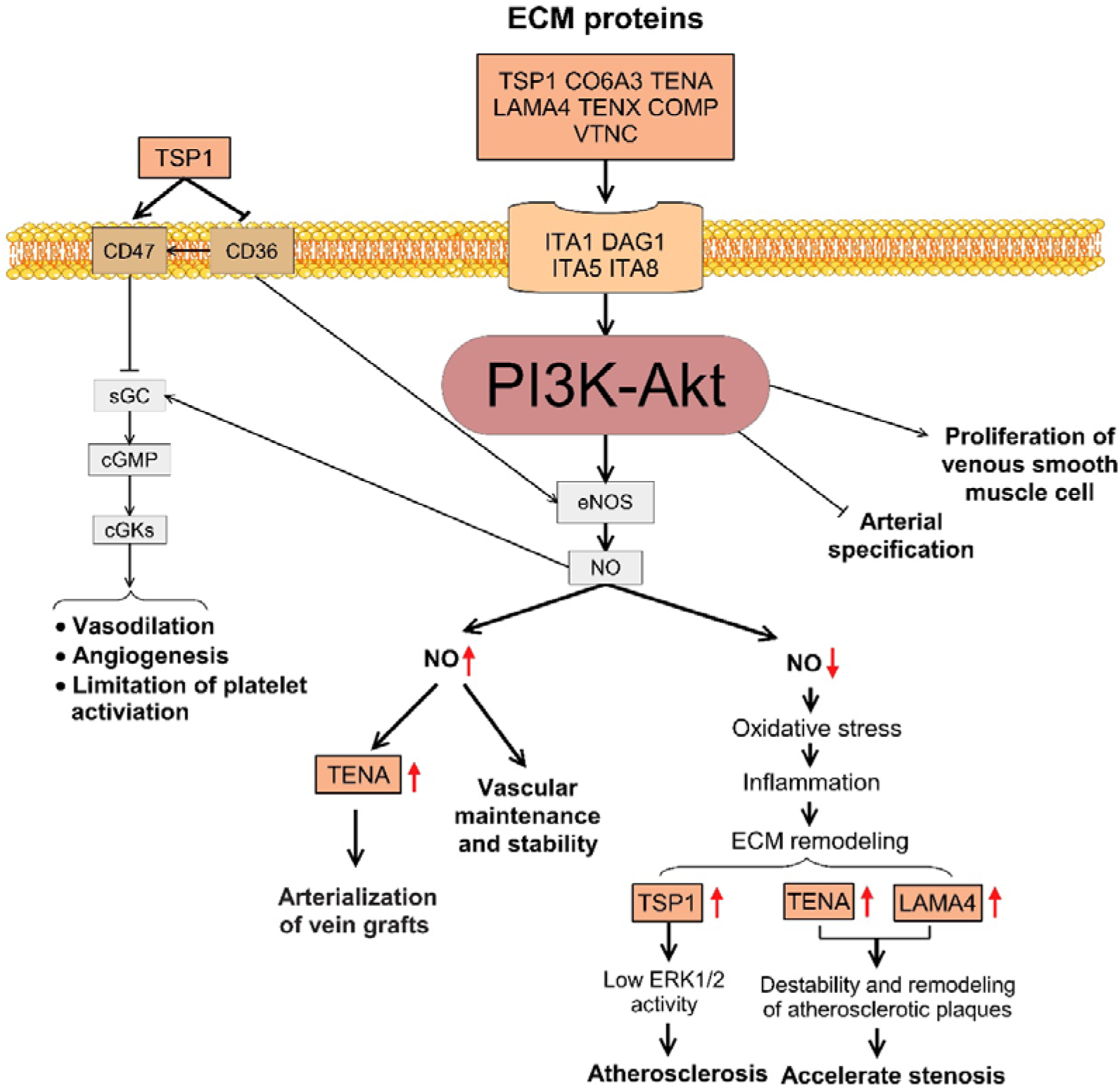
ECM-PI3K-Akt pathway interacted with NO among the three grafts. A proposed model for the central role of ECM-PI3K-Akt pathway interacted with NO-cGMP pathway. TSP1, CO6A3, TENA, LAMA4, TENX, and COMP interacted with receptors (ITA1, CD44, DAG5, ITA5, and ITA8) are significantly enriched in the ECM and PI3K-Akt pathway. All these proteins, particularly TSP1, TENA, and LAMA4, have certain connections with nitric oxide and play important roles in vasodilation, stenosis, angiogenesis, platelet activation, inflammation, ECM remolding, and atherosclerosis. All those mechanisms are related to long-term patency. IMA, internal mammary artery; RA, radial artery; SV, saphenous vein; ECM, extracellular matrix; PI3K, phosphatidylinositol-4, 5-bisphosphate 3-kinase; Akt, protein kinase B; eNOS, endothelial nitric oxide synthase; NO, nitric oxide; sGC, soluble guanylyl cyclase; cGMP, cyclic guanosine monophosphate; cGKs, cGMP-dependent protein kinases.

### Differences between the arteries and the vein

Major differences were found between the artery (IMA or RA) and vein (**Fig. 5B**). All these pathways may play important roles in atherosclerosis. p53 exerts critical regulatory effects in atherosclerosis^20^ whereas deacetylated p53 plays a pro-survival role in oxygenated myocardial infarction hearts by binding endothelial nitric oxide synthase gene promoter^20^. Interestingly, TSP1 can be inhibited by the inactivation of p53^21^. The lower expression of TSP1 in the arteries than in the vein may explain why arteries have less atherosclerosis and higher patency.

### Significance of lower TSP1 in IMA or RA than in SV

Increase of NO production is mediated by PI3K-Akt pathway activation^22, 23^. In contrast, NO signaling can be inhibited by TSP1 in vascular smooth muscle cells^24^, platelets^25^, and endothelial cells by binding two of receptors, CD36 and CD47^26^. Lower expression of TSP1 in both IMA and RA than SV found in this study indicates that arteries may potentially release more NO than the vein as reported by us^10^ before.

TSP1 facilitates the growth and migration of vascular smooth muscle cells^27^. Antibody blockade of TSP1 accelerates reendothelialization and reduces neointima formation in rat carotid artery^27^. In addition, high level of TSP1 inhibits ERK1/2 activity and results in atherosclerosis^28^. Taken together, the lower level of TSP1 in arteries than in veins found in this study likely correlates to least the better long-term patency of IMA than that of venous grafts.

### Patency of the arterial grafts (IMA and RA) is higher than SV grafts

The present study is not a clinical follow-up study; it is a biological experimental study with graft samples from patients who simultaneously received IMA, RA, and SV grafts and had redundant graft tissue available for experiments. Those conditions limited the number of the patients in this study. Even in this limited number of patients with limited time of follow-up, the patency was significantly different between the arterial grafts (IMA and RA) and SV. In fact, all the 28 arterial grafts were patent in comparison with significant stenosis appeared in 4 of the 19 SV grafts (p<0.05; **Table S17**).

### TENA is lower in IMA than that in SV or RA

The expression of TENA (Tenascin-C) is associated with intimal hyperplasia and atherosclerosis^29^. TENA expression declined in situations where intimal hyperplasia is inhibited, such as prostaglandin E_2_ deficiency^30^ or treatment with a NO donor^31^. TENA is a crucial molecule in neointimal hyperplasia of artery grafts at an early stage and can accelerate artery graft stenosis, which is the reason why TENA is recognized as a prime target for therapy to inhibit smooth muscle cell migration and proliferation in free artery grafts^32^. Lower expression of TENA was found in IMA than that in RA and SV and this may be involved in higher long-term patency of IMA compared to RA or SV.

### Differences between the two arteries

We also found differences between the two arteries. Among the 5 DE mRNAs and proteins, ITA8 and AL1A1 were only different in the comparison between the two arteries (**Fig. 5B** and **Fig. 6F**). The atherosclerotic lesions can depress the activity of ALDH1 (coding gene of AL1A1) and eventually favor oxidative stress and the severity of atherosclerosis^33^. The higher expression of ALDH1 in IMA than in RA may indicate why IMA have less atherosclerosis and higher patency than RA.

## Limitations

This study was performed in trios of the major CABG grafts (IMA, RA, and SV) from the patients simultaneously receiving these three grafts, so the samples are limited. Further animal studies will be performed to clarify the mechanisms between the biological differences and the long-term patency using the DE RNAs and DEPs.

## Conclusions

In summary, this study demonstrated the biological differences among three CABG grafting vessels as conduits in 4 levels (non-DE RNAs/non-DEPs, DE RNAs/non-DEPs, non-DE RNAs/DEPs, and DE RNAs/DEPs) by using transcriptomics and proteomics. ECM-PI3K-Akt pathway with correlated DE mRNAs and proteins (TSP1, TENA, TENX, VTNC, LAMA4, CO6A3, COMP, ITA1, DAG1, ITA5, and ITA8) was found to have major differences in the development of stenosis, angiogenesis, and atherosclerosis. This study provides new insights into the answer for why the grafts have different patency at the biological basis and into the new therapeutic targets for improving the results of CABG.

## Data availability

The data underlying this article will be shared on reasonable request to the corresponding author.

## Acknowledgements

The authors thank all the nurses for the assistance of collecting grafts, Department of Cardiovascular Surgery, TEDA International Cardiovascular Hospital, Tianjin, China.

## Author Contribution

GWH conceived the whole study. HTH performed experiments. HTH and GWH wrote the manuscript. HTH, HXC, ZQW, and JW collected the samples. HTH, HXC, JW, and QY prepared materials for experiments. QY discussed the study protocols, the process of the experiments, and the manuscript. GWH had overall responsibility for the whole study. All authors approved the final manuscript version.

## Ethical Approval

The protocol was approved by the Ethics Committee of the TEDA International Cardiovascular Hospital ([2019]-0826-4). Informed consent was obtained before surgery. All samples were collected according to the principles outlined in the Declaration of Helsinki.

## Nonstandard Abbreviations and Acronyms

CABG: coronary artery bypass grafting
cGMP: cyclic guanosine monophosphate
DE: differentially expressed
DEPs: differentially expressed proteins
eNOS: endothelial nitric oxide synthase
FPKM: Fragments Per Kilobase Million
GO: Gene Ontology
IMA: internal mammary artery
KEGG: Kyoto Encyclopedia of Genes and Genomes
MRM: Multiple Reaction Monitoring
NO: nitric oxide
RA: radial artery
SV: saphenous vein

