## Supplemental figures and tables for "Integrative Multi-omics Approach Reveals the Molecular Characterization and Differences of ECM-PI3K-Akt Pathway among Coronary Artery Bypass Grafting Conduits with Clinical Implications": Hou He- IMA RA SV-Supplementary Materials-2024.8.2.docx

August 1, 2024

**with Clinical Implications**

**Running Title: Artery/Vein Grafts in Coronary Surgery**

**Hai-Tao Hou^1,2^, Huan-Xin Chen^1,2^, Zheng-Qing Wang^1,2^, Lei Xi^1,2^, Jun Wang^1,2^, Qin Yang^1,2^, Guo-Wei He^1,2*^**

^1^Department of Cardiovascular Surgery & Institute of Cardiovascular Diseases, TEDA International Cardiovascular Hospital, Chinese Academy of Medical Sciences & Peking Union Medical College & Tianjin University, Tianjin, China

^2^Tianjin Key Laboratory of Molecular Regulation of Cardiovascular Diseases and Translational Medicine, Tianjin, China

^*^**Correspondence author:**

**Professor Guo-Wei He, MD, PhD, DSc**

**Distinguished Professor of Tianjin University**

**Foreign Correspondence Member, The National Academy of Medicine, France.**

**&**

**Clinical Professor of Surgery, Oregon Health and Science University, Portland, Oregon, U.S.A.**

TEDA International Cardiovascular Hospital, No.61, the 3rd Ave, TEDA, Tianjin, 300457

**This file includes** **Supporting Information**

**Supplementary Figure S1.** Correlation analyses of transcriptomics and proteomics.

**Supplementary Figure S2.** Enriched GO categories of mRNAs, proteins, and correlations in three grafts.

**Supplementary Table S1**. Differentially expressed mRNAs in IMA-SV (in separate excel file).

**Supplementary Table S2**. Differentially expressed lncRNAs in IMA-SV (in separate excel file).

**Supplementary Table S3**. Differentially expressed circRNAs in IMA-SV (in separate excel file).

**Supplementary Table S4**. Differentially expressed mRNAs in RA-SV (in separate excel file).

**Supplementary Table S5**. Differentially expressed lncRNAs in RA-SV (in separate excel file).

**Supplementary Table S6**. Differentially expressed circRNAs in RA-SV (in separate excel file).

**Supplementary Table S7**. Differentially expressed mRNAs in IMA-RA (in separate excel file).

**Supplementary Table S8**. Differentially expressed lncRNAs in IMA-RA (in separate excel file).

**Supplementary Table S9**. Differentially expressed circRNAs in IMA-RA (in separate excel file).

**Supplementary Table S10.** Sequences of primer for qRT-PCR.

**Supplementary Table S11**. Differentially expressed proteins in IMA-SV (in separate excel file).

**Supplementary Table S12**. Differentially expressed proteins in RA-SV (in separate excel file).

**Supplementary Table S13**. Differentially expressed proteins in IMA-RA (in separate excel file).

**Supplementary Table S14**. Correlations between RNA and protein expression in IMA-SV (in separate excel file).

**Supplementary Table S15**. Correlations between RNA and protein expression in RA-SV (in separate excel file).

**Supplementary Table S16**. Correlations between RNA and protein expression in IMA-RA (in separate excel file).

**Supplementary Table S17**. Patency of the grafts from 13 patients. Note there was no stenosis in either IMA or RA.

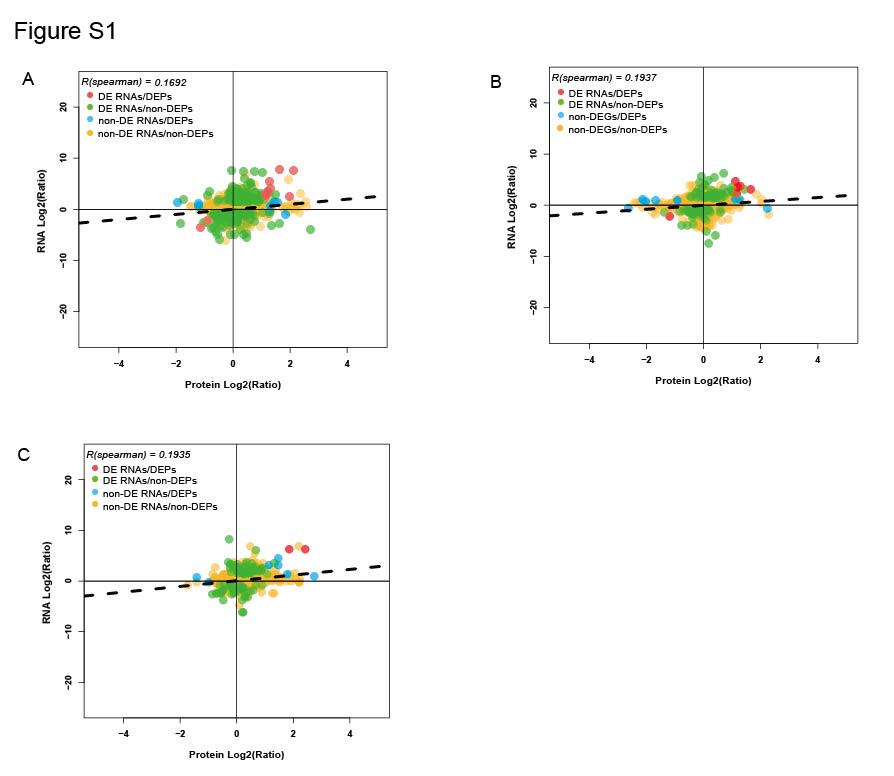

Figure S1. **Correlation analyses of transcriptomics and proteomics.**

Red points: DE RNAs and DEPs; blue points: DE RNAs and non-DEPs; green points: non-DE RNAs and DEPs; yellow points: non-DE RNAs and non-DEPs.

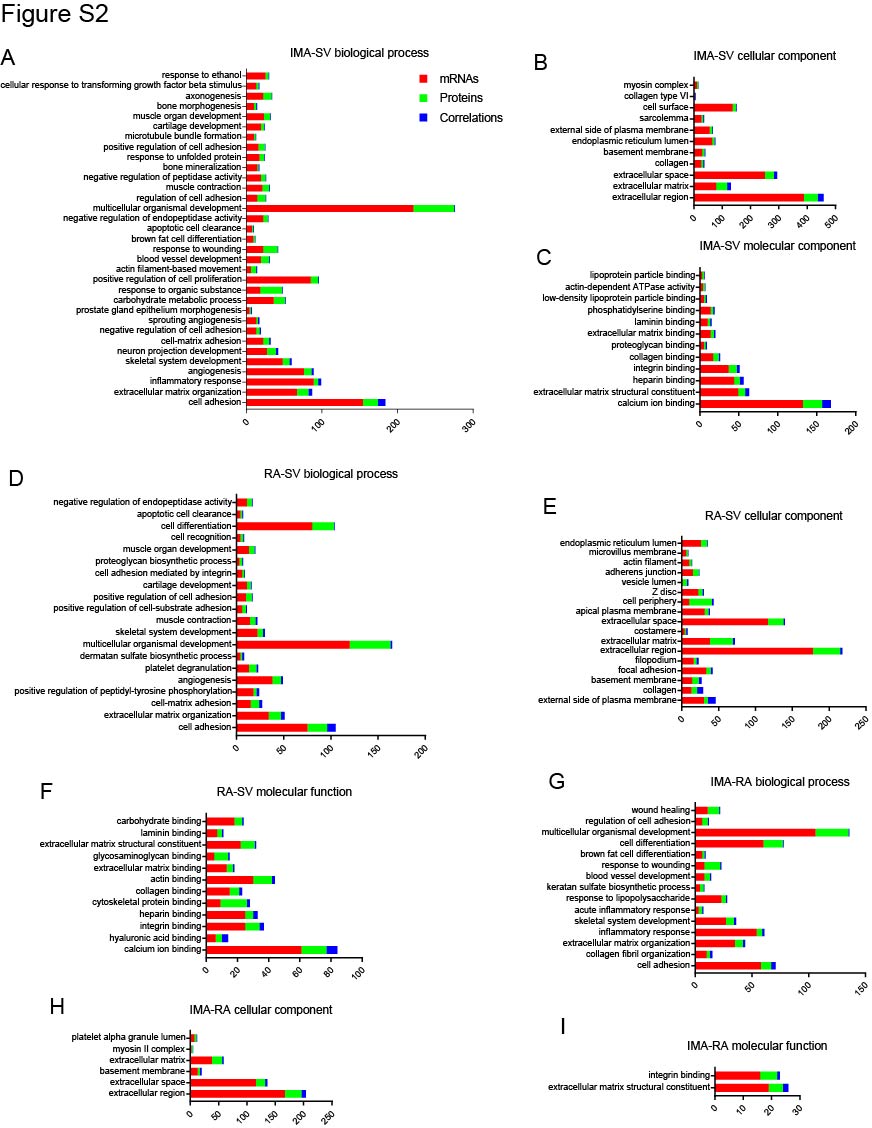

Figure S2. **Enriched GO categories of mRNAs, proteins, and correlations in 3 grafts.**

Correlations analyses about biological process, cellular compartment, and molecular function using differentially expressed mRNAs and differentially expressed proteins in IMA-SV (A-C), RA-SV (D-F), and IMA-RA (G-I).

**Table S10.** Sequences of primer for qRT-PCR

| **Gene** | **Forward primer** | **Reverse primer** |
| --- | --- | --- |
| MYH10 | GATGAGAAGCGGCGTCTGGAAG | CTTGCGGAAGCGGTCGTTGAG |
| GJA3 | CACCACCACCTGCTGATGACTG | AGACTGCTGCCGCTCCCATC |
| FBP2 | TCCATCGGAACCATCTTTGCCATC | ACGCCTTGCCCTGTGGAGAG |
| CCL7 | GCACTTCTGTGTCTGCTGCTCAC | AGTGGCTACTGGTGGTCCTTCTG |
| VMP1 | GATTGAAGCCTGCATGTGG | TCTGGTTCAGCACCTGAGAG |
| ITLN1 | TGCTGGAATGAGGGTCACCG | CGGCTGCTGCTGTAACCAAC |
| PCK1 | AGGCGGCTGAAGAAGTATGACAAC | CCCTGGGAACCTGGCATTGAAC |
| DES | TCAATGACCGCTTCGCCAACTAC | CCCGCAGCTCCTCCTCGTAG |
| GJA5 | GGAGGAAGGGAATGGAAGGATTGC | CATGGTGGTGCGGATCAGGATG |
| S100A9 | GAACACATCATGGAGGACCTGGAC | GGTTAGCCTCGCCATCAGCATG |
| MSX2 | CCTTTACCACATCCCAGCTC | ACTCTGCACGCTCTGCAAT |
| HOXC10 | ATCAAGACGGAGCAGAGCCT | GCACCTCTTCTTCCTTCCGC |
| PRAC2 | ATGGACAGAAGGCGGATGGC | CCAGGAAGAAGGCAAGGAGGT |
| HOXC13 | CGCGGCTAGCAAGTTCATCA | AGGTGGAGTGGAGATGAGGC |
| HMCN2 | GCTGCTCAGCCTCTGGATAC | CTGTGCGTCCACATGGATT |
| THBS1 | TACGCTGGCAATGGCATCATCTG | GCACACAAAGGACCTGGCTCTAC |
| HOXC11 | AACAAGAACAGCGTCCTGCC | TGTTCTCCTCCTCAGCCTCC |
| KRTAP1-5 | TGCGGATTTCCCAGCTTCTCAAC | ATGCCACCACCAATGCCACAG |
| TNC | TCCATCATCCTGACACCACCTCTC | GCACCTCCTCCATAGCAACTAAGC |
| NRP2 | GCCCTCGAAGGTCTCTCACT | TGTTCTGGGCCTCGAAATGC |
| GAPDH | AACAGCGACACCCACTCCTC | GGAGGGGAGATTCAGTGTGGT |

**Table S17.** Patency of the grafts from 13 patients. Note there was no stenosis in either IMA or RA.

|  | IMA | RA | vein |
| --- | --- | --- | --- |
| Patient Number | 13 | | |
| Graft Number | 13 | 15 | 19 |
|  | 28 (IMA+RA) | | 19 |
| Graft with Stenosis | 0 | | 4 |
| *P* value | 0.022 | | |
